# Trends and Factors associated with Neonatal Mortality among Neonates Hospitalized at the Women and Newborn Hospital in Lusaka Zambia

**DOI:** 10.1101/2023.07.26.23293218

**Authors:** Deborah Tembo, Patrick Kaonga, Cheelo Mweene, Choolwe Jacobs

## Abstract

**Background:** Neonatal mortality is a global problem especially in resource-poor settings such as sub-Saharan African Countries. Trends and factors associated with neonatal mortality vary in different settings. This cross-sectional study investigated trends and factors that are associated with increased neonatal mortality among hospitalised neonates with a view of identifying pointers that can be modified to help improve neonatal survival.

**Methods:** Medical records for all neonates admitted to the NICU in 2018 and 2019 were reviewed for the study. The Mortality rates were calculated using WHO standard and Microsoft excel 2010 used to construct monthly trends analysis using monthly totals. Univariate and multivariate logistic regression analysis to determine the factors associated with neonatal deaths at the tertiary facility using Stata version 14.2. Model fit was evaluated using Hosmer and Lemeshow test (chi2=13.90, P =0.0845), implies the model’s estimates fit the data at an acceptable level.

**Results:** A total of 7,581 admissions were seen and 2340 files were extracted for analysis, of which 940 (40.2%) died. The overall Neonatal mortality percentage during the study period was 31.8%. Relatively similar trends were observed during the study period with differences in case fatality rates. Factors associated with an increased odds of dying were not attending antenatal care (AOR=2.09, 95% CI [1.46 - 2.99] p <0.0001), parity (AOR=1.09, 95% CI [1.02 – 1.16] p=0.0013) and age of the neonate in days (AOR=0.92, 95% CI [0.91- 0.94] p <0.0001).

**Conclusion:** Neonatal mortality is high among hospitalized neonates at the Women and New-born Hospital in Zambia. Associated Factors included no antenatal attendance, increased parity, and early age of the neonate. This has detrimental effects; hence emphasis is on early Antenatal Care for easy monitoring of mother and neonates for fast actions in case of complications

## INTRODUCTION

Neonatal mortality is a global problem especially in resource-poor settings. It is a major contributor to under five mortalities. Globally estimated at 2.7 million deaths yearly [1, 2]. The WHO (2018) estimates that in Africa, an average of 15% of newborn babies are expected to die before the age 28 days. The corresponding figures for many parts of the world which are developed such Europe, and USA are much lower ranging from 2 to 3% or even less [1]. Neonatal mortality rates in sub-Saharan Africa was estimated at 27 deaths per 1000 live births in 2017 [1] . The Zambia demographic health survey estimates neonatal mortality rate at 27 deaths per 1000 live births in 2018 [3] , this is unacceptably high. Literatures shows that the trends and factors associated with the Neonatal mortalities vary in different settings and mostly not known. Causes of neonatal deaths are largely preventable and treatable using proven, cost-effective and currently available interventions [4]. Studies have shown that, a complex chain of factors are associated with neonatal deaths ranging from socio-economic, biological to healthcare-related factors [5, 6]. According to a study by Qazi and Stoll [7], Three conditions: infection, birth asphyxia, and consequences of premature birth/low birth weight, are responsible for majority of these deaths [7].

In Zambia, many strategies are being implemented to help reduce neonatal deaths and improve neonatal survival. Programs such as helping baby breath campaign, emergency obstetric and neonatal care training of health workers (EmONC) [8], are being implemented to help achieve legacy goal No 1 of 2020 to reduce neonatal mortality rate to12 per 1000 live birth. Moreover, the Zambia University teaching hospital (UTH) in 2017 was divided into 5 hospitals which included the women and newborn hospital to decentralize the management and improve client care. However, there is limited evidence of studies conducted at facilities using routinely collected hospital data to examine the impact these strategies in improving newborn survival. Yet analysis of routine data is useful in monitoring performance and progress made towards reducing neonatal deaths.

Hence this study sought to investigate the trends and factors associated with neonatal mortality among neonates admitted at the Neonatal Intensive care Unit Women and Newborn Hospital with a view of identifying pointers that can be modified to help reduce the current Mortality levels

### Objectives

The study tried to estimate the neonatal mortality rates, assess the monthly trends in neonatal deaths and determine the associated factors in neonatal deaths at NICU women and newborn hospital UTH over the past two years, Zambia between 2018 to 2019

## Methods

### Study Design and setting

This study was a retrospective cross-sectional study. Data for all the neonatal admissions reported in the hospital data bases and case files were extracted at NICU from January 2018 to December 2019. The study was conducted at the neonatal intensive care unit (NICU)-women and newborn hospital, University teaching hospital, Zambia. It is the largest tertiary hospital and main referral health institution in Zambia located approximately 4km to the east of the city Centre in the capital city Lusaka.

### Study Participants

The study population were newborn babies admitted at Neonatal Intensive Care Unit (both discharges and deaths) between 2018 January to December 2019. All admissions of neonates that were made within first 28 days of life were included in the sample. Case files for admitted neonates were used to collect the variables used in the study however files with missing and incomplete data were in the sample was excluded from the study. The sample was collected using Total enumeration method. All routinely monthly aggregated data for admissions, discharges and deaths found at the Unit from January 2018 to December 2019 was included in the analysis of the trends and mortality rate.

### Variables

The study outcomes were to estimate neonatal mortality rates, assess monthly trends in neonatal deaths and determine the associated factors in neonatal deaths at NICU women and newborn hospital while independent variables including age of the neonate, cause of death, place of delivery, gestational age, mode of delivery. residence, mothers age, parity, maternal HIV status

### Study size

A Complete enumeration of all reported cases of neonates admitted at NICU-women and newborn hospital-UTH (all cases in the period under review) during the period of January 2018 to December 2019 were used. Through the case files data was extracted for both alive and died. However, sample size calculation done manually using sample size for prevalence. The minimum sample was 360 with the size power of about 80%, however a complete enumeration was used to improve substantial power to above 80% which increases the accuracy of the predictions. Total complete files extracted from the two-year admissions were 2340. A data collection tool with variables needed in the study was used to collect the data from patient files, registers, and monthly departmental reports (HMIS)

### Sources of data

The source of the data were the medical files, registers, and reports at Neonatal Intensive Care Unit department. The data was obtained in a process which began from registry to the ward. Patient medical files were incorporated in the Health Management Information Systems records and the data collected in weekly reporting tools then later aggregated in the monthly and quarterly Health Management Information System tools. The case files contained information on the neonate’s individual characteristics including age, sex, diagnosis, and mode of separation i.e., 18 alive or died. The clerk’s data submits reports to the Hospital information systems officer who are the custodians of case files. 3.8 Data extraction and tools Data was extracted from all reports and case files found at Neonatal Intensive Care Unit data department from January 2018 to December 2019. The causes of deaths were analyzed with respect to clinical information in the files. There was an ongoing multidisciplinary consultation with the information managers, ward in charges and staffs at different levels at Neonatal Intensive Care Unit to verify the data. A data extraction tool was developed to extract relevant information from the data bases. To Ensure validity and reliability data from the medical files and registers both admissions and discharges were compared. The data extracted was used to identify key characteristics of all the neonates used as participants in this study. Data was then coded entered and stored in Microsoft excel then exported to Stata for analysis.

### Ethical considerations

Permission to use data collected routinely at NICU was sought from the Ministry of Health for analysis. Confidentiality was maintained as this data was restricted. A waiver of consent and ethical approval was gotten from the University of Zambia Biomedical Research Ethics Committee (Ref. No 244 2019) as there was no contact with the actual individuals. Permission was also sought from the Zambia national research authority who approved the research to be conducted. The data was stored as encrypted files on all computers to prevent access, accidental loss, or destruction. The data will be kept safe for as long as it is necessary for this research purpose only **Quantitative variables**

### Statistical Methods

The data collection tools were used to collect all the data needed. Data stored in excel then cleaned and exported to Stata.The data was then checked for completeness and accuracy by ensuring that, the information answers and addresses all the questions in the data collection tool. Responses were categorized using statistical measuring scales, coded, and entered on the data master sheet for easy analysis Data was destringed, entered and analysed in STATA version 14.2 College Station, Texas 77845 USA.

We used world health organization (WHO) mortality rate calculation, i.e., the number of deaths divided by the total number of live admissions per 1000. Monthly trends were assessed using Microsoft excel. Preliminary data analysis involved description of predictor variables to understand their distribution in relation to dependent variable (Neonatal death) through tabulations. In descriptive statistics, median and Interquartile range were reported for Age of the neonate, Gestation at Birth, Birth weight, maternal age, Number of pregnancies and parity. Shapiro-Wilk W test for normal data test was used to test the data for skewness of all continuous variables and count variables. chi-square test was used to check for associations on all categorical data The Man-Whitney test was done to check for comparison differences in continuous data and chi-square test was used to check for associations on all categorical data.

Neonatal mortality rate among the neonates admitted to NICU was calculated using WHO standard of calculation, that is the number of deaths during a certain time interval divided by total number of live births per 1000, the live births in this study were the neonates who were admitted at the unit and excel was used to check the trends over the years in months assuming all the assumptions for the model were met. Both univariate and multivariable logistic regression were done to describe the relationship between the dependent variable ‘new-born deaths’ with the selected predictor variables with adjusted odds ratios (AOR) at 95% confidence intervals (CI). An investigator lead backwards regression was used according to literature on neonatal deaths and goodness of fit was used to check for the best fit model.

## Results

### Participants/ Descriptive data

During the study period a total of 7581 neonates were hospitalised at NICU with different conditions ranging from infections to congenital disabilities. Among the admissions 5171 were discharged alive while 2410 died. Of those 5241 files were excluded from analysis of the factors associated to neonatal mortality because they had incomplete charts and some charts were not accessed due to various reasons. We managed to extract 2340 complete charts or files for analysis, 1400 (59.83%) neonates were discharged alive and 940 (40.17%) died. Of those 1,223 (52,26 %) were males and 1117 (47.74%) were females. The median age of the neonates was 4 days (IQR 2- 7) while the median gestational age was 36 weeks (IQR 30 - 37). The median birth weight was 2400g (IQR 1500 - 3000). The median maternal age was 25yrs (IQR 21 - 31) while median number of pregnancies for the mothers was 2 (IQR 1 - 4) and the median maternal parity was 2 (IQR 1 - 3). Among the neonates, 2134 (91.07%) were delivered at the hospital, 104 (4.44%) health centres and 105(4.49%) at home as shown in Table 1

**Table 1.** Basic Characteristics of The Mother and Neonates.

| Variable |  | Median/IQR |
| --- | --- | --- |
| Age (days)* |  | 1 (2, 7) |
| Gestational age (weeks)* |  | 36 (30, 37) |
| Age of Mother(years)* |  | 25 (21, 31) |
| The number of pregnancies* |  | 2 (1, 4) |
| Number of children* |  | 2(1, 3) |
| Category |  | Frequency (%) |
| Sex | Male | 1,223 (52.26%) |
|  | Female | 1,117 (47.74%) |
| Place of Birth | Hospital | 2,134 (91.07%) |
|  | Heath centre | 104 (4.44%) |
|  | Home | 105(4.49%) |
| Status | Alive | 1,400 (59.83%) |
|  | Died | 940 (40.17%) |
| Type of Birth | Single | 2,235 (95.51%) |
|  | Twin | 105 (4.49%) |
| Mode of Delivery | Assisted Breech Delivery | 50 (2.14%) |
|  | Caesarean- Section | 328 (14.02%) |
|  | Spontaneous Vaginal Delivery | 1,949 (83.29%) |
|  | Vacuum-assisted Vaginal Delivery | 13 (0.56%) |
| Maternal HIV Status | Known Negative | 1,784 (76.24) |
|  | Known Positive | 387 (16.54 %) |
|  | Unknown | 169(7.22%) |
| Attended Antenatal Care | No | 220 (9.40%) |
|  | Unknown | 154 (6.58%) |
|  | Yes | 1,966 (84.02%) |
| Condition | Mono – condition | 598(25.6%) |
|  | Co-conditions | 1742(74.4%) |

\*The median and Interquartile range were reported. Shapiro-Wilk W test for normal data test was used to test the skewness of all continuous variables and count variables

And analysis of the age using a box plot (figure 1) showed that there was increased survival in older neonates as compared to those with fewer days. The median age for neonates that were alive was 5 days (IQR 3 - 8) and the those that died was 2 days (IQR 2- 5) with the p-value

**Figure 1.**
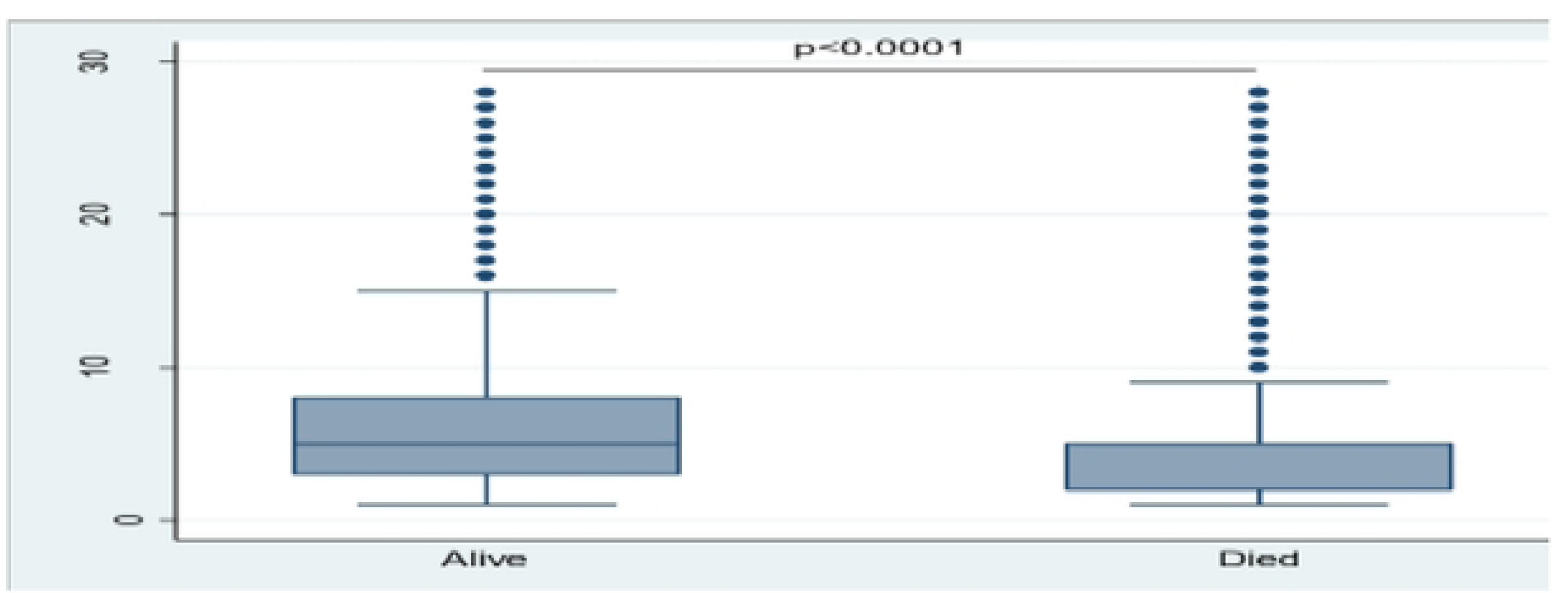
Box plot of Association between Age of neonates that were discharge alive and those that were discharged dead.

### Outcome Data

#### Neonatal Mortality rate among the admitted Neonates

The overall percentage of neonatal mortality among neonates admitted to NICU Women and New-born hospital for the two years (2018,2019) was 31.8 % as shown in the table 2. The results show evidence of a differences in the neonatal mortality rates between the two years. 2019 had the highest mortality percentages of 36.6% while 2018 had 27.2 %.

**Table 2:** Crude Neonatal mortality rates for NICU-mother and new-born hospital-UTH (2018-2019)

| Year | Discharged alive |  | Died |  |  |
| --- | --- | --- | --- | --- | --- |
|  | Total discharges | Percentage (%) | Total deaths | Rate per 1000 admissions | Percentage (%) |
| 2018<br>(N=3867) | 2815 | 72.8% | 1,052 | 272 | 27.2% |
| 2019<br>(N=3714) | 2356 | 63.4% | 1,358 | 366 | 36.6% |
| Cumulative<br>(N=7581) | 5171 | 68.2% | 2,410 | 318 | 31.8% |

**P-value <0.0001**\*these calculations are based on institutional data only for the neonates admitted to NICU for the two years. The calculation was done using the world health standard of number of deaths divided by the total number of live births for the given period of time per 1000. And the live births in this situation were the total number of live admissions.

**Table 3:** univariate and multivariate logistic regression analysis of the outcome (Died) with independent variables.

| Predictor | CRUDE |  |  | ADJUSTED |  |  |
| --- | --- | --- | --- | --- | --- | --- |
|  | OR* | p-value | 95%CI* | OR* | p-value | 95%CI* |
| **Age Neonate** | 0.92 | <0.001 | (0.90, 0.94) | 0.92 | <0.001 | (0.90,0.94) |
| **Sex** |  |  |  |  |  |  |
| Male | Ref |  |  |  |  |  |
| Female | 1.20 | 0.030 | (1.02, 1.42) | 0.99 | 0.854 | (0.78,1.23) |
| **Gestation age at birth** |  |  |  |  |  |  |
|  | 0.82 | <0.001 | (0.81, 0.84) | 0.94 | 0.004 | (0.90,0.98) |
| **Birthweight** | 1.00 | <0.001 | (1.00, 1.00) | 1.00 | <0.001 | (1.00, 1.00) |
| **No of pregnancies** | 1.02 | 0.446 | (0.97,1.073) | 0.80 | 0.022 | (0.66, 0.97) |
| **No of Children** | 1.06 | 0.042 | (1.00, 1.11) | 1.27 | 0.016 | (1.05,1.55) |
| **HIV status** |  |  |  |  |  |  |
| Negative | Ref |  |  |  |  |  |
| Positive | 1.44 | <0.001 | (1.56, 1.80) | 0.96 | 0.803 | (0.79, 1.31) |
| Unknown | 1.19 | 0.277 | (0.87, 1.64) | 1.15 | 0.613 | (0.69,1.87) |
| **Place of Birth \*\*** |  |  |  |  |  |  |
| Hospital | Ref |  |  |  |  |  |
| Health centre | 2.06 | <0.001 | (1.38, 3.08) | 2.66 | <0.001 | (1.55 .4.57) |
| Home | 3.35 | <0.001 | (2.20, 5.10) | 2.43 | 0.979 | 0 |
| **Mode of Delivery\*\*** |  |  |  |  |  |  |
| SVD | Ref |  |  |  |  |  |
| C-Section | 1.83 | <0.001 | (1.45, 2.31) | 1.74 | 0.659 | (0.15,20.44) |
| Breech-VD | 15.28 | <0.001 | (6.03,38.7) | 5.56 | 0.002 | (1.83,16.54) |
| Vacuum-VD | 20.83 | 0.004 | (2.70, 160.5) | 64.36 | 0.010 | (2.71, 1528.11) |
| **Multiple Birth** |  |  |  |  |  |  |
| No | Ref |  |  |  |  |  |
| Yes | 3.28 | <0.001 | (2.16, 5.00) | 0.99 | 0.983 | (0.57, 1.74) |
| **Delivered by Midwife** | Ref |  |  |  |  |  |
| Doctor | 1.98 | 0.001 | (1.58, 2.49) | 2.33 | 0.497 | (0.20,26.86) |
| Unskilled | 3.47 | 0.001 | (2.28, 5.26) | 0.00 | 0.981 | 0 |
| **Antenatal Attendance** |  |  |  |  |  |  |
| Yes | Ref |  |  |  |  |  |
| No | 2.73 | <0.0001 | 2.04, 3.65) | 2.35 | <0.0001 | (1.56, 3.55) |
| Unknown | 0.39 | <0.0001 | (0.26, 0.59) | 0.19 | <0.0001 | (0.12, 0.33) |

\*OR- odds ratio, CI -confidence interval
\*\* some variables had wider confidence interval but was checked for variance inflation factor and multiple collinearity and could be due to big differences in variable frequencies

#### Trends in hospital Neonatal mortality

The trends in neonatal mortality for the two years were similar but there was increased case fatality for 2019 with a slightly high numbers of neonatal deaths compared to 2018. The numbers were almost the same for July 2019 and July 2018 as shown in figure 2.

**Figure 2.**
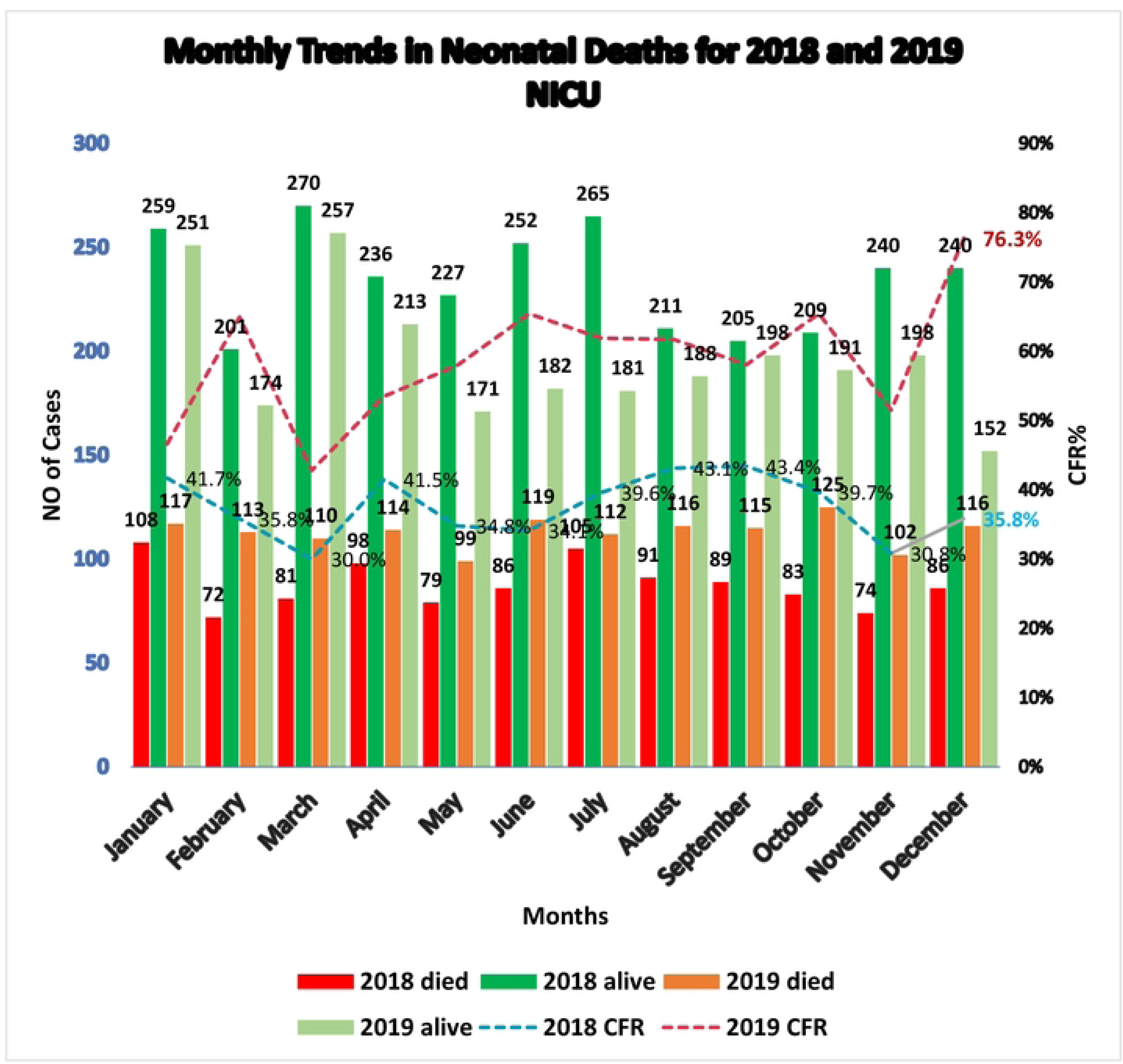
Monthly trends in neonatal deaths for 2018 and 2019.

#### Factors associated with Neonatal Mortality at the Neonatal Intensive Care Unit (NICU)

When univariate logistic regression analysis was conducted, an increase in the age of the neonate(days), had 8% reduction in dying (COR= 0.92, 95% CI [ 0.92,0.94], p<0.0001) while the increase in the number of children increased the risk of neonatal deaths by 94% (COR=1.06, 95% CI [1.00,1.11] p= 0.042). Interestingly the gestational age was associated with reduced odds of new-born deaths (COR= 0.82,95% CI [1.02,1,42], p<0.0001). Prematurity as a primary diagnosis compared to birth asphyxia increased the odds of new-born death 3 times (97%), (COR= 3.03, 95% CI [ 2.38,3.87], p<0.001. Birthweight, positive HIV status, place of birth, mode of delivery and some residences were found to increase the odds of neonatal deaths.

At multivariate logistic regression one-day increase in the age of the neonate was associated with reduced odds by about 8% (AOR=0.92, 95% CI [ 0.92,0.94], p<0.0001) likelihood of neonatal death compared to a smaller number of days. An increase in the number of pregnancies was associated with 20% reduced odds of new-born deaths (AOR:0.80,95% CI [ 0.66,0.98] p = 0.022) and 73% the increase in the number of children was associated with an increased odds of new- born deaths (AOR= 1.27 95% CI [1.04,1,55], p=0.016). Gestational age, Birthweight, place of birth, mode of delivery and some residences were found to associated neonatal deaths

A model for best predictors shows that one-day increase in age of the neonate predicted less chance of neonatal deaths (AOR=0.92, 95% CI [0.91- 0.94] p <0.0001) taking account of gestational age, parity, antenatal attendance, place of birth, mode of delivery and primary diagnosis. A unity increases in parity had an increased chance of having neonatal death by 1.09 which could be as low as 1.02 and as high as 1.16. (p= 0.0013), controlling for all variables in the model. Mothers who did not attend antenatal care had an increased chance of having their neonates dying compared to those who attended antenatal services (AOR=2.09, 95% CI [1.46 – 2.99] p<0.001), controlling for all variables in the model. Delivering from home, complicated delivery i.e., C-section vacuum or breech all increased the chances of having neonatal deaths controlling for all variables in the model. 1-unit increase in gestational age had less chance of having neonatal death considering all the variables in the model as shown in table 4

**Table 4.**
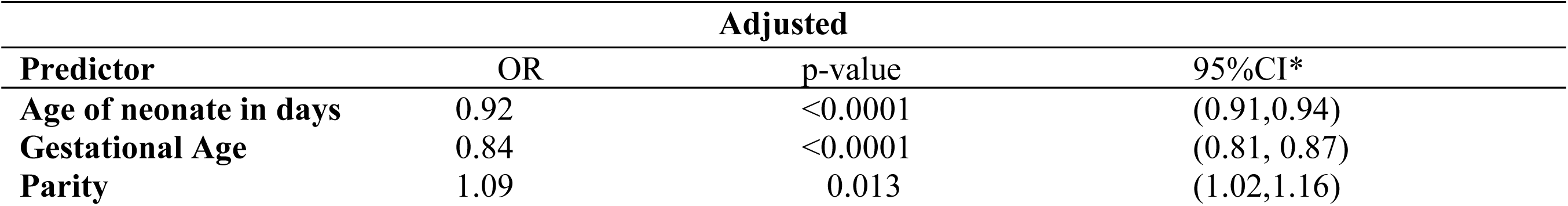
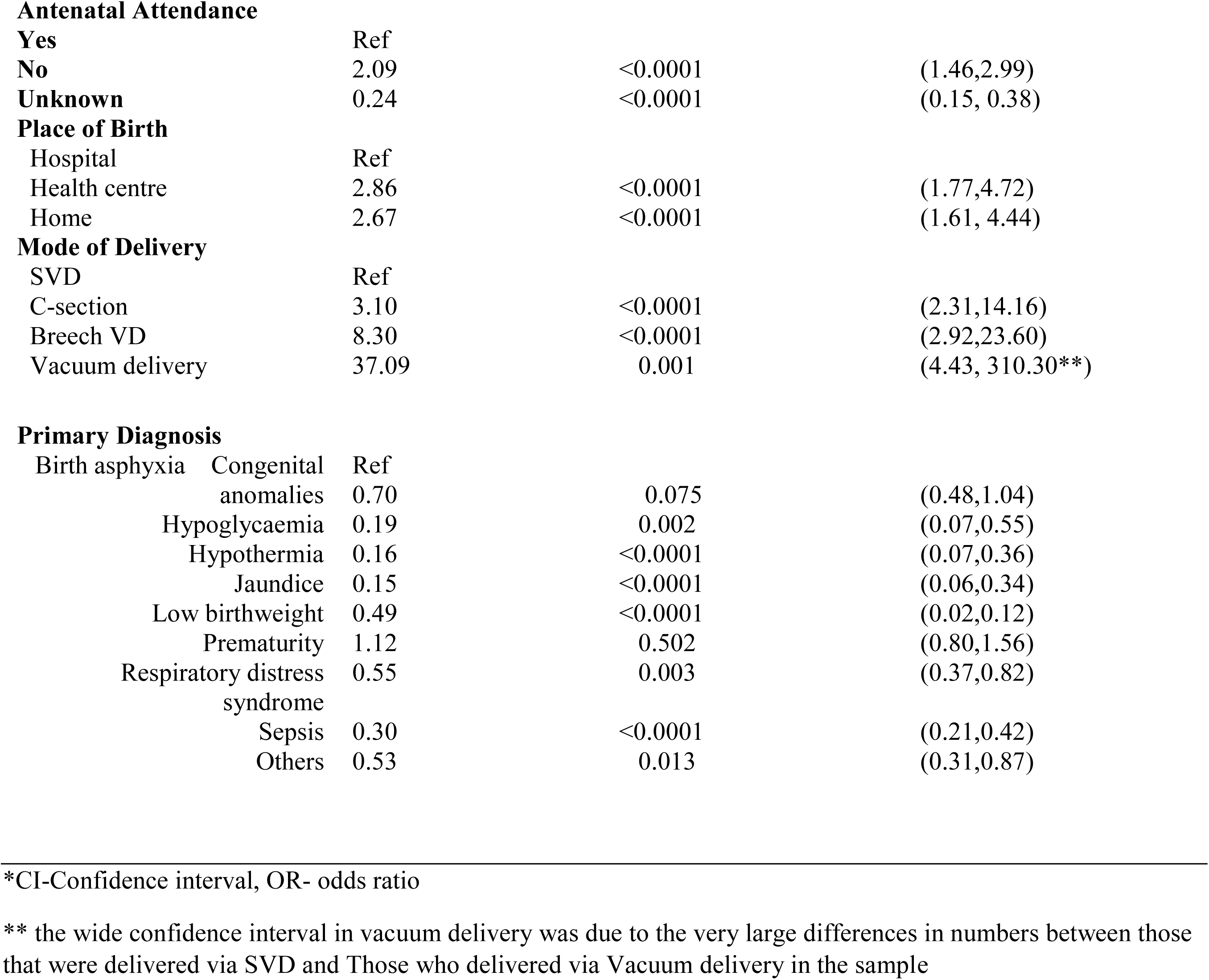
The best predictors model logistic regression for factors associated with new-born deaths.

## DISCUSSION

The study aimed at investigating the trends and factors that are associated with increased neonatal mortality amongst neonates admitted to Neonatal Intensive Care Unit Women and Newborn Hospital, UTH Zambia over the past two years (2018-2019). In this study Neonatal mortality among the admission was significantly high with the overall neonatal mortality among the admitted neonates was higher than the findings of a Ghanaian study that had an overall neonatal mortality at 20.2% [9]. The reason could be due to the high levels of prematurity which was 26.6 % of all the deaths. In a hospital-based study conducted in Ethiopia , the hospital Neonatal Mortality was 5.7% which was much lower than our findings [10]. Similarly , mortality percentages were still lower than the findings in the current study in studies conducted in Ghana 16.0% [11], northern Nigeria 16.9% [12], Ethiopia at University of Gonder 14.3% , Felege hiwot referral hospital 13.29% and India 11.41% [13–15] . This is unacceptably high and might be regarded as one of the contributors to the country’s high neonatal mortality rate that showed a 3% increase from 24 deaths per 1000 live per for 2013-2014 Demographic Health Survey to 27 per 1000 live births for the 2018 Demographic Health Survey. World Health Organization targets to reduce neonatal mortality to as low as 12 deaths per 1000 live births according to the Sustainable Development Goals of 2030 targets. Although different researches like that by Hadgu, the lancet collaborators and Owusu have documented evidence that show higher mortality percentages in male than female neonates this study found no significant evidence of the differences, this could be attributed to large differences in samples sizes between the two sexes [9, 16, 17].

Taking a glimpse at the graphs of the trend data, it revealed a relatively similar trend in neonatal deaths over the two years. But the case fatality rate was higher for 2019 ending at 76.3% in December 2019 compared to 35.8% for December 2018. This implies an upward adjustment in trends in neonatal deaths for the facility. This increase in the trends might be due to an increase in neonatal admission for the year 2019 or the severity of the cases like increase in prematurity. According to a Tanzanian study the Overall, the neonatal mortality trend demonstrated an apparent stagnation during the 10-year period, with similar patterns observed in 2006–2010 and 2011–2015 despite interventions instituted during the early 2010s [18]. The findings in this study indicated clearly that the number of hospital neonatal deaths continued unabated despite the implementation of various interventions like helping the baby breath, centralized management, and trainings for newborn care [8, 19]. In another study the findings indicated no significant decline was observed in neonatal mortality trends in the study period [20]. These signifies the need to move our attention to ways to improve the interventions to achieve the desired pattern of improved trends in neonatal deaths in hospitals which could spill over to help improve neonatal survival and overall attain neonatal mortality goals. In another study at a neonatal intensive care unit in Tehran, Iran, they found that mortality rate was decreasing year by year (Jaberi Zahra *et al*.,2013). Hence there is urgent need for the responsible health personnel to share notes with such facilities to help find lasting solutions to health in attaining the much-needed reduction in neonatal mortalities.

The study also showed that there are several factors that play a role in the huge crude neonatal mortality rates although it is understood that the neonates admitted to the specialized facilities are already having major challenges. The factors associated to neonatal mortality at NICU women and newborn hospital found in this study included: early neonatal age, premature labour, or low gestational age, increase in parity, birthweight, mode of delivery like vacuum deliveries, place of delivery like home, who assisted in the delivery like unskilled birth attendants and diagnoses like prematurity, birth asphyxia and sepsis. These finding were like a study in Zambia by Lukonga were common causes of neonatal mortality in Zambia were sepsis, birth asphyxia, and prematurity/low birth weight and most of these deaths occur within the first 24 hours of life [5]. It seems that the interventions such emergency obstetric care (EMOC), helping the baby breath and other related health messages being implemented over the years have not done much in reducing the burden of neonatal mortality at the highest referral in Zambia.

## Conclusion

The study findings have revealed the need to evaluate the strategies that are currently being implemented to reduce neonatal mortality. There is urgent need to re-strategize if we are to attain the targets of the sustainable development goals of 2030.The 2018–2019 Neonatal intensive care data at University Teaching Hospital, Women and New-born Hospital has demonstrated no significant changes in neonatal mortality trend. Early Neonatal age, low gestational age, increased parity, not attending antenatal care and prematurity were found to be main predictors of neonatal mortality. These findings need appropriate attention for care of women in childbearing before pregnancy, during pregnancy and after delivery to enable prompt care in cases of high risky conditions. Institutional delivery should be promoted and strengthened to reduce home delivery. The findings in this study provides substantial evidence for the care providers; program implementers and policymakers and help them to pass evidence-based,decisions and take timely interventions within the facilities of neonatal intensive care.

### Limitations

**S**econdary data from the HMIS was used hence there was inconsistencies, incompleteness, missing information which paused as a challenge to the research. The study does not permit one to draw causal association between neonatal deaths and the associated factors. However, the Scientific evidence provides milestones on neonatal health which may be important to measuring progress made in achieving neonatal mortality goals.

## Data Availability

The authors here by declare that all the data for this study are available for access in case of Validation, replication, reanalysis, new analysis, reinterpretation or inclusion into meta-analyses and Reproducibility of research. The data can be access through the university of Zambia school of public health Library and portal. Also, the co- author can be reached at Gmail –, Phone +260977998260. All citations have been added to the manuscript

## Abbreviations

CSO: Central Statistics Office
MDG: Millennium Developmental Goals
MoH: Ministry of Health
MPDSR: Maternal Prenatal Death Surveillance and Response
HMIS: Health Management Information System
NICU: Neonatal Intensive Care Unit
NMR: Neonatal Mortality Rate
SSA: Sub-Saharan Africa
SDGs: Sustainable Developmental Goals
UNICEF: United Nations Children Fund
UTH: University Teaching Hospital

## Contributions

DT designed the study, performed the statistical analysis and data interpretation, and wrote the manuscript, methodology. PK, CJ contributed to the study design, analysis, interpretation of and supervision of the study. All authors reviewed and approved the final Manuscript

## Availability of data and materials

Data was easily accessible at the University Teaching Hospital Lusaka Zambia, Mother, and newborn hospital.

## Acknowledgement

The authors appreciated the assistance of the staff of Ministry of health M and E department, Lusaka, Zambia regarding sampling/listing procedures. University teaching Hospital management for allowing Us to conduct the study at the facility, Staff for Neonatal Intensive Care Unit women and new-born UTH for their various tasks and the support during data collection.

## Summary Box

### What is already known on this topic

Studies that have been conducted in Zambia have shown a high Neonatal mortality, rates at 27 /1000 live births according to the 2018 Zambia demographic health survey. The factors that contribute to these deaths are complex and include but not limited to socio-economic, biological, and healthcare-related factors. However, there was limited evidence of studies conducted at facility level (hospital level) using routinely collected hospital data to examine the neonatal mortality rates and contributions to the high neonatal mortality rates at facility level.

### What this study adds

This study has provided evidence on the most recent trends in neonatal mortality and has bridged the knowledge gap on associated factors to neonatal deaths at NICU- women and newborn hospital Zambia.

### How this study might affect research, practice, or policy

The Scientific evidence that have been provided stimulates the need to intensify monitoring of pregnancy from inception to delivery and up to 6 weeks after to ensure appropriate neonatal care. This also implores policy- makers and implementers on the need to re-design and re-strategize current interventions and policies on promoting good quality maternal health before and during pregnancy, child birth and after delivery so as to improve neonatal survival..

